# Ambient scribe in general practice: a multi-perspective before-after longitudinal mixed-methods study

**DOI:** 10.1101/2025.10.30.25339135

**Authors:** R.C.A. van Linschoten, C.M. van Loon, L. Joanknecht, E.W.M.A. Bischoff

## Abstract

**Background:** High workload among general practitioners (GPs) poses a growing challenge to clinician well-being and care quality. Ambient scribes offer a potential solution, but evidence on their effectiveness is limited and largely overlooks the patient perspective.

**Methods:** A prospective multicentre multi-perspective before-after longitudinal mixed-methods study with 12 GPs and GPs in training without prior experience using ambient scribes from the Netherlands. The intervention was an ambient scribe, which generates summaries of consultations using a microphone and large language model. The primary objective was to evaluate the effect on clinical documentation time. Secondary outcomes included total consultation time, documentation quantity and quality, patient and GP experiences, tool acceptability, and usage rate. Outcomes were observed across a two-day baseline and two day intervention period. Quantitative data were analysed using (generalized) linear mixed models, and interviews were thematically analysed. This trial was registered at ClinicalTrials.gov, NCT06691724.

**Findings:** Between December 9, 2024, and July 2, 2025, 12 GPs and 535 patients (264 baseline, 271 intervention) were included in the study. Clinical documentation time decreased by 42.7 seconds (95% CI [-56.29; - 30.78], p<0.0001). No difference was found in total consultation time (-61.4 seconds, 95% CI: –131.91; 0.96; p=0.069). GPs reported reduced workload, and some patients reported improved patient-provider communication. However, potential drawbacks were inaccurate summaries, barriers for discussing sensitive topics, and possible interference with the clinician’s reasoning process.

**Interpretation:** Ambient scribing decreases workload and may improve communication. However, potential unintended consequences need further investigation to ensure that care quality and accessibility are not affected.

**Funding:** Rijnmond Dokters and the Erasmus Trustfund.

## Introduction

Increasing workload in healthcare is a major global concern, affecting clinicians across specialties and settings.(1, 2) General practitioners (GPs) are particularly vulnerable to rising work demands.(3) In the Netherlands, 68% of GPs report their workload as too high, with workload increasing over the years, and nearly one in five describe their work as very or extremely stressful.(3) This chronic strain has resulted in significant staffing challenges: over half of Dutch GP practices struggle to recruit locum GPs or support staff, and 60% have stopped accepting new patients.(4-6) High workload negatively affects both provider well-being and the quality of care delivered, impacting job satisfaction, patient-provider communication, healthcare costs, care quality, and patient safety.(7-9)

A major contributor to workload is the administrative burden.(3) GPs in the Netherlands spend only 60% of their time on direct patient care, with the remaining 40% devoted to indirect or non-patient-related activities, such as documentation and administrative tasks.(6) Increasing time spent on clinical documentation is linked to higher stress levels and reduced time for patient interaction, key drivers of physician dissatisfaction and burnout.(4, 10, 11)

Reducing the time GPs spend on documentation has the potential to improve physician well-being and patient care. Recently, ambient scribes have emerged as a promising tool. These systems use large language models (LLMs) to passively record and transcribe medical consultations in real-time, generating clinical documentation with minimal physician input. This would allow physicians to focus on patient interaction while the system handles documentation in the background.

Research on ambient scribes has shown some promising benefits for clinical practice. Retrospective before-and-after studies using electronic health record (EHR) log data suggest that ambient scribes may reduce documentation time.(12-16) However, these studies are not controlled and may be subject to bias. The only study using the gold standard of continuous external observation found no significant change in documentation time when using an ambient scribe.(17, 18) Similarly, a different study evaluating productivity also reported no change when an ambient scribe was introduced.(19)

Most evaluations of ambient scribes have reported reduced perceived workload and cognitive burden among clinicians,(12, 14-16, 20-24) though some studies found no effect.(18, 25) Evidence regarding communication and patient experience is mixed: while some studies report improved communication,(14, 21-24) others found no change.(12, 19) Importantly, almost all research on communication has focused on the clinician perspective. Only one study included the patient experience data via a survey, and none employed qualitative methods to explore the patient experience in depth.

Despite growing interest in ambient scribes, existing evidence is limited in both scope and methodological quality. Most studies use retrospective designs and rely on indirect measures, with little attention for the patient perspective. Consequently, the impact of ambient scribes on efficiency and care quality remains insufficiently understood. To address these gaps, we conducted a study to comprehensively evaluate the use of ambient scribes in clinical practice from both the provider and patient perspectives.

## Methods

### Study design

This prospective multicentre multi-perspective longitudinal before-after mixed-methods study examined the effect of ambient scribe in general practice. This study was conducted in general practices across the Netherlands between December 2024 and July 2025. The medical ethical review committee of the Erasmus MC deemed the study to not be subject to the Medical Research Involving Human Subjects Act and provided a waiver (MEC-2024-0286).

### Participants

The study included two participant groups: GPs or GPs in training interested in implementing the ambient scribe and patients who had consultations with these GPs. GPs were recruited through regional care groups in the Netherlands and the professional networks of the authors. GPs were purposively selected to ensure diversity in both patient populations (e.g., age, migration background, gender) and GP demographics, with the goal of obtaining a representative and generalizable sample. GPs with prior experience using AI-based transcription and reporting tools during consultations were excluded from participation.

### Intervention

The intervention under study was the Juvoly QuickConsult transcription tool.(26) This software records and transcribes consultations using a microphone and an LLM. Hereafter, an LLM generates a structured summary of the consultation based on the Subjective, Objective, Assessment, and Plan (SOAP) framework. SOAP is the standard format for clinical documentation in general practice. In some EHRs, Juvoly QuickConsult is directly integrated. When integration was unavailable, GPs used the website from Juvoly QuickConsult and manually copied and pasted the generated summary into their EHR.

### Timing

The study was conducted across three consecutive phases: a baseline period, an implementation period, and an intervention period, each lasting two weeks. During the baseline period, GPs were observed over two full days or three half-days of consultations without using the ambient scribe. During the implementation period, GPs received a brief explanation and demonstration of the tool, followed by time to practice using it. During the intervention period, GPs were again observed for two full days or three half-days while using the ambient scribe.

### Outcomes

The primary outcome was the time spent on clinical documentation in seconds, which included time waiting for the tool to generate summaries as well as for copying and pasting the summaries into the EHR if applicable. Secondary outcomes included total time per consultation in seconds, the quantity and quality of documentation, GP and patient experiences, the acceptability of the tool, and the tool usage rate. For training and validity of outcome assessors, see the Supplementary Methods. Investigators, patients, and providers were not blinded to intervention allocation as this was not feasible.

During both the baseline and intervention period, time outcomes were assessed through direct observation of GP consultations. An external observer continuously recorded the time spent on different tasks using specialized software.(27) Tasks were categorized as either communication-related or hands-on; definitions for each task type are provided in Supplementary Table 1. Multitasking was calculated by measuring the overlap between communication and hands-on tasks.

Documentation quantity was reported as the number of words used in each SOAP-item. Documentation quality was assessed by counting the number of relevant clinical variables in the note, which were categorized by type: signs and related variables (e.g. site, onset, character, time course), contextual factors (e.g. self-care, risk behaviour, family history), symptoms and measurements (e.g. physical examination, pulse, temperature), (differential) diagnosis or plan variables (e.g. prescriptions, referrals, safety net).

Patient experiences were assessed during both the baseline and intervention period using the Patient Experience Questionnaire (PEQ).(28) This survey asked patients to evaluate their experiences during the consultation with the GP across different categories: medical content, communication, general experiences, and their emotions immediately after the consultation. Scores for medical content, communication, and general experiences ranged from 1 (indicating a negative outcome) to 5 (indicating a positive outcome), while emotions were rated on a scale from 1 (negative emotion) to 7 (positive emotion).

To gain deeper insight into the experiences of both patients and GPs with the ambient scribe and its impact on the consultation, semi-structured interviews were conducted during the intervention period. These interviews were guided by a predefined topic list (see Supplementary Methods). Interviews were conducted face-to-face with all participating GPs. Patients were approached after consultations and asked whether they would be willing to participate in an interview. Those who agreed received an information letter about the study, and invited for an interview. Patient interviews were conducted face-to-face, via Microsoft Teams, or by phone, preferably within the same week as the consultation. Each interview lasted around 10-20 minutes and was audio-recorded and transcribed using Microsoft Teams. Transcripts were not returned to patients or GPs for comments or corrections.

The acceptability and usage of the tool by GPs were assessed during the baseline, implementation, and intervention period using the Unified Theory of Acceptance and Use of Technology (UTAUT) questionnaire.(29) The questionnaire covers outcome expectation, effort expectation, social influence, facilitating conditions, and intention to use the tool. Scores ranged from 1 (negative outcome) to 5 (positive outcome).

For each GP included in the study, the following baseline characteristics were collected: gender, age, years of experience as a GP, type of practice, patient population size, percentage of patients from deprived areas, and whether the ambient scribe was directly integrated into the EHR. Digital skills (categorized in general digital skills, EHR skills, skills for safe internet use, and competencies using various programs and applications) were assessed using the Dutch digital skill questionnaire for general practice.(30) The geographic area of the practice was defined according to the criteria of the Dutch Central Bureau of Statistics (CBS).(31)

For each patient observed in the study, age and gender were collected. We also collected the International Classification of Primary Care (ICPC) codes linked to the consultation from the EHR.(32) For each patient interviewed, we also collected gender, educational level, migration background, language proficiency, and presence of long-term health conditions.

### Sample size

Sample size calculations were based on simulations that explored various assumptions to assess their impact on the required sample size. The assumptions included:

- Average consultation duration of 10 or 15 minutes, with a standard deviation (SD) of 5 minutes. (33, 34)
- Average documentation time equal to 35% of the total consultation time (SD 5%), as well as a more conservative estimate of 17.5% (SD = 2.5%).(35, 36)
- A normally distributed random effect at the GP level to account for variation in documentation time, with an SD of 1 or 2 minutes.
- A minimum documentation time of 10 seconds per consultation.
- A reduction in documentation time of either 25% or 50% due to the use of the tool.

Even under the most conservative assumptions: a documentation time of 1.75 minutes, a GP level random effect of 2 and an effect size of 0.75, 10 consultations each for 3 GPs would provide a statistical power of 99%.

To enhance the generalizability of our findings, we aimed to include at least ten GPs from at least five different practices. For each GP, we observed consultations over two full days or three half-days, yielding approximately 40 consultations per GP and an expected total of 400 observations per study period. The sample size for interviews varied: we interviewed all participating GPs and all patients who expressed willingness to be interviewed.

### Statistical analysis

We conducted an intention-to-treat analysis comparing consultations during the intervention period with those from the baseline period. All data were analysed as observed. The impact of the ambient scribe on clinical documentation time and total consultation time was assessed using linear mixed models. These models accounted for clustering by including a random effect for each participating GP and a fixed effect for planned consultation duration.

To evaluate documentation quantity and quality, we used negative binomial generalized linear mixed models with a log link function, appropriate for the distribution of the outcome variables. These models also included a random effect for participating GPs and a fixed effect for planned consultation time. Patient experience questionnaires were analysed using linear mixed models with random effects for participating GPs. All random effects were assumed to follow a normal distribution. Tool usage rates and acceptability were summarized using descriptive statistics.

Three sensitivity analyses were performed. First, a per-protocol analysis was conducted for time, documentation and patient experience outcomes. In the per-protocol analysis, we included consultations from the baseline period only when the tool was not used, and from the intervention period only when the tool was used. Second, we included an additional adjustment for total non-documentation time when analysing documentation time and documentation-related outcomes. This variable was considered a potential confounder for two reasons. Within the fixed 10- or 15-minute consultation window, reduced patient-facing time may increase the time available for documentation. Moreover, more extensive in-consultation discussions may lead to increased documentation. Third, because there were differences in the reasons for consultations between the baseline and intervention period, a post-hoc sensitivity analysis was conducted for time outcomes which we adjusted for ICPC code.

All quantitative analyses were conducted using R (version 4.4.3) within the R-studio environment (version 2025.05.0). Mixed models were fitted using lme4, lmerTest was used for p-values and 95% bootstrap confidence intervals (CI), and model diagnostics were assessed using the DHARMa package.(37-39)

### Qualitative analysis

Semi-structured interview transcripts were coded by the second author (CvL) using MaxQDA (version 24.7.0) and analysed using thematic analysis with an inductive approach. The process began with open coding, followed by axial coding to group codes into themes and categories, helping to identify patterns within the data. Analysis was conducted in multiple stages to uncover recurring themes across interviews, with regular discussions among the research team to refine the interpretation and structure of codes and themes. To ensure consistency, the first author (RvL) independently coded two transcripts using the final code tree, and any discrepancies were resolved through discussion.

## Results

Between December 9, 2024, and July 2, 2025, twelve GPs participated in the study, contributing a total of 535 observed consultations (Table 1; see also Figure 1 and Supplementary Table 2). Most consultations addressed a single complaint, though some involved multiple issues. In total, 333 complaints were recorded in 264 consultations during the baseline period and 339 complaints in 271 consultations during the intervention period.

**Table 1.** Baseline characteristics of GPs.

| Characteristic<sup>1</sup> | N = 12 |
| --- | --- |
| GP characteristics |  |
| Age (years) | 39 (37, 49) |
| Female | 8 (67%) |
| Experience as a GP (years) | 8 (5, 15) |
| Digital skills |  |
| General digital skills | 9.30 (8.20, 9.90) |
| EHR skills | 7.60 (5.15, 8.40) |
| Skills in using various programs | 8.60 (6.85, 9.45) |
| Skills for safe internet use | 7.80 (7.35, 8.75) |
| UTAUT |  |
| Outcome expectation | 4.00 (3.25, 4.75) |
| Effort expectation | 4.00 (3.75, 4.75) |
| Social influence | 3.38 (2.88, 3.63) |
| Facilitating conditions | 3.50 (3.25, 4.00) |
| Intention to use | 4.00 (3.33, 4.33) |
| Practice characteristics |  |
| Type |  |
| Duo practice | 3 (25%) |
| Group practice | 9 (75%) |
| List size | 5,791 (4,733, 6,585) |
| Location |  |
| Rural | 3 (25%) |
| Semi-rural | 2 (17%) |
| Urban | 7 (58%) |
| Patients from a deprived area | 2% (2%, 4%) |
| Direct link between tool and EHR | 2 (17%) |
| <sup>1</sup> Median (Q1, Q3); n (%);<br>Abbreviations: EHR=electronic health record, GP=general practitioner,<br>UTAUT=Unified Theory of Acceptance and Use of Technology<br>**Table 1.** Baseline characteristics of GPs |  |

**Figure 1.**
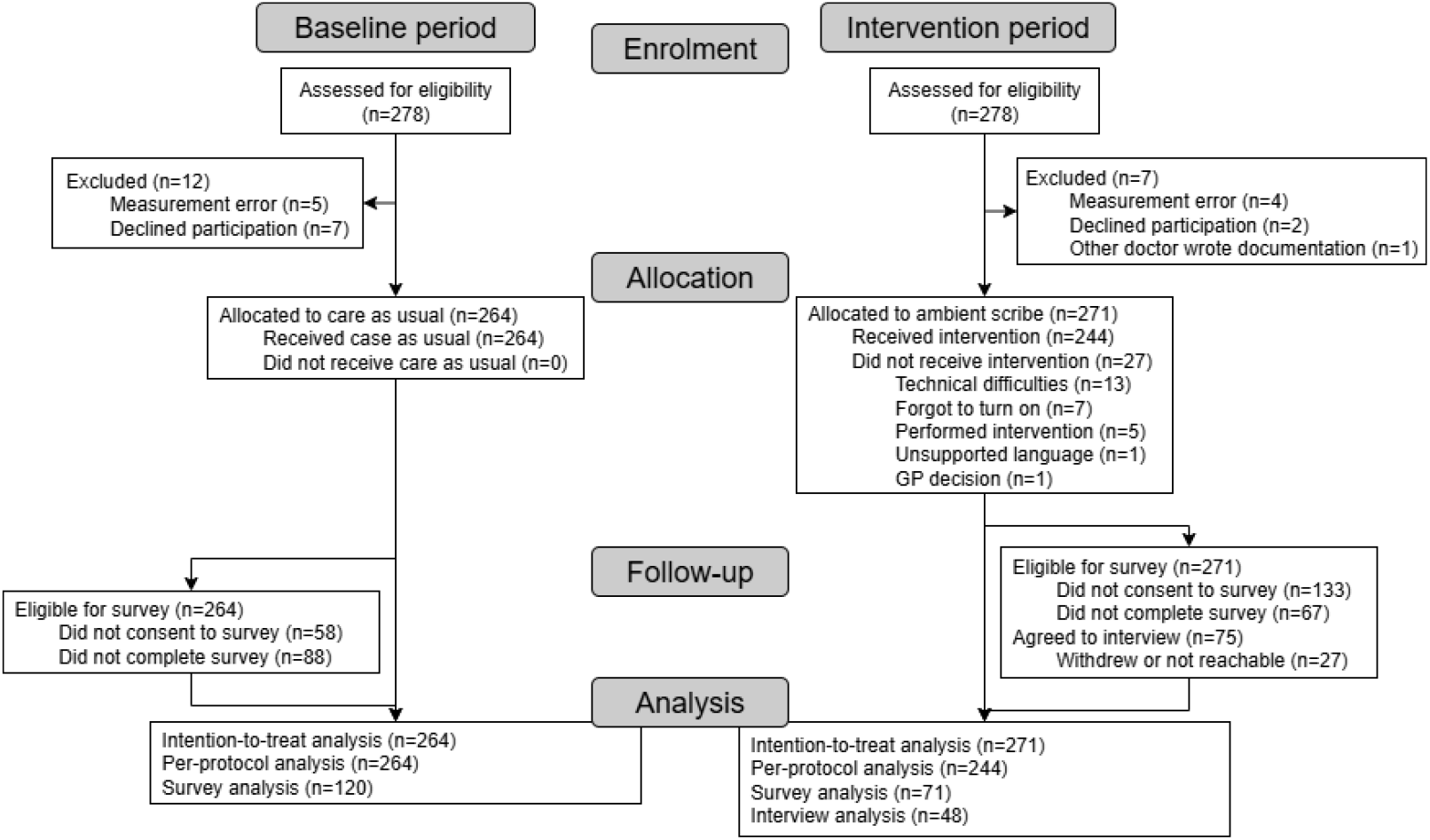
Flowchart of patient inclusion

The majority of consultations were scheduled for 15 minutes, 91% during the baseline period and 89% during the intervention period. General and unspecified complaints were more frequent in the baseline period (absolute percentage difference: +4.6%), along with ear issues (+2.5%). In contrast, complaints related to the nervous system (+3.2%) and skin (+2%) were more commonly discussed during the intervention period (Supplementary Table 3). Missing data were minimal for most variables, except for survey responses (see Table 1 and Supplementary Tables 2 and 3).

### Primary outcome

During the intervention period, the mean time spent on documentation per consultation was 42.7 seconds shorter than in the baseline period (95% CI: -56.29 to -30.78; p < 0.0001).

### Secondary outcomes

The total consultation time showed no significant difference (-61.4 seconds, 95% CI: –131.91 to 0.96; p = 0.069). For distribution of time variables between the baseline and intervention period, see Supplementary Table 4. There was an increase in the length of the subjective, assessment and plan parts of the clinical note in the intervention group, but not in the objective part. In the intervention period, there were more sign and plan variables noted in the documentation, but less symptom and measurement variables, and no significant difference was found in the number of context and diagnosis variables. No differences were found in patient experience as measured with the patient experience questionnaire between the intervention and baseline period. See Table 2 for all estimates, confidence intervals and p-values.

**Table 2.** Parameter estimates, 95% CIs and p-values of the main analyses.

| Name | Estimate | 95% CI | p-value |
| --- | --- | --- | --- |
| **Time outcomes<sup>1</sup>** |  |  |  |
| Documentation time | -42.72 | [-56.29; -30.78] | <0.0001 |
| Consultation time | -61.38 | [-131.91; 0.96] | 0.069 |
| **Note length<sup>2</sup>** |  |  |  |
| Subjective | 1.15 | [1.04; 1.27] | 0.005 |
| Objective | 0.87 | [0.73; 1.04] | 0.131 |
| Assessment | 1.19 | [1.01; 1.4] | 0.036 |
| Plan | 1.57 | [1.41; 1.75] | <0.0001 |
| **Number of variables in note<sup>2</sup>** |  |  |  |
| Signs | 1.12 | [1.01; 1.22] | 0.036 |
| Context | 1.03 | [0.89; 1.18] | 0.621 |
| Symptoms | 0.80 | [0.69; 0.92] | 0.004 |
| Diagnosis | 0.97 | [0.84; 1.14] | 0.713 |
| Plan | 1.18 | [1.08; 1.29] | 0.0004 |
| **Patient experience<sup>1</sup>** |  |  |  |
| Outcome of the consultation | 0.10 | [-0.14; 0.34] | 0.427 |
| Communication experiences | 0.11 | [-0.09; 0.33] | 0.286 |
| Absence of communication barriers | -0.03 | [-0.22; 0.17] | 0.773 |
| Staff relationship | 0.03 | [-0.16; 0.23] | 0.770 |
| Emotions after consultation | 0.08 | [-0.28; 0.45] | 0.682 |
| Abbreviations: CI=confidence interval |  |  |  |
| <sup>1</sup> Difference on an additive scale |  |  |  |
| <sup>2</sup> Difference on a multiplicative scale |  |  |  |
| **Table 2.** Parameter estimates, 95% CIs and p-values of the main analyses |  |  |  |

The sensitivity analyses adjusted for total documentation time showed the same results (Supplementary Table 5). The only difference between the per-protocol analysis and intention-to-treat analyses was that in the per-protocol analysis the number of context variables in the note was higher in the intervention period than in the baseline period (Supplementary Table 6). The post-hoc sensitivity analysis adjusted for ICPC code agreed with the main analyses (Supplementary Table 7).

Nearly all patients in the intervention group were invited to participate in interviews, with a few exceptions due to logistical issues, emergency consultations, or a language barrier. Of the 75 patients who initially agreed, 48 were interviewed (Figure 1). Participant demographics are presented in Supplementary Table 8. No new themes emerged in the final interviews, suggesting that data saturation was reached. Thematic analysis identified six overarching themes: process of using the tool, communication, clinical documentation, the consultation, safety of the tool, and workload (Table 3).

**Table 3.**
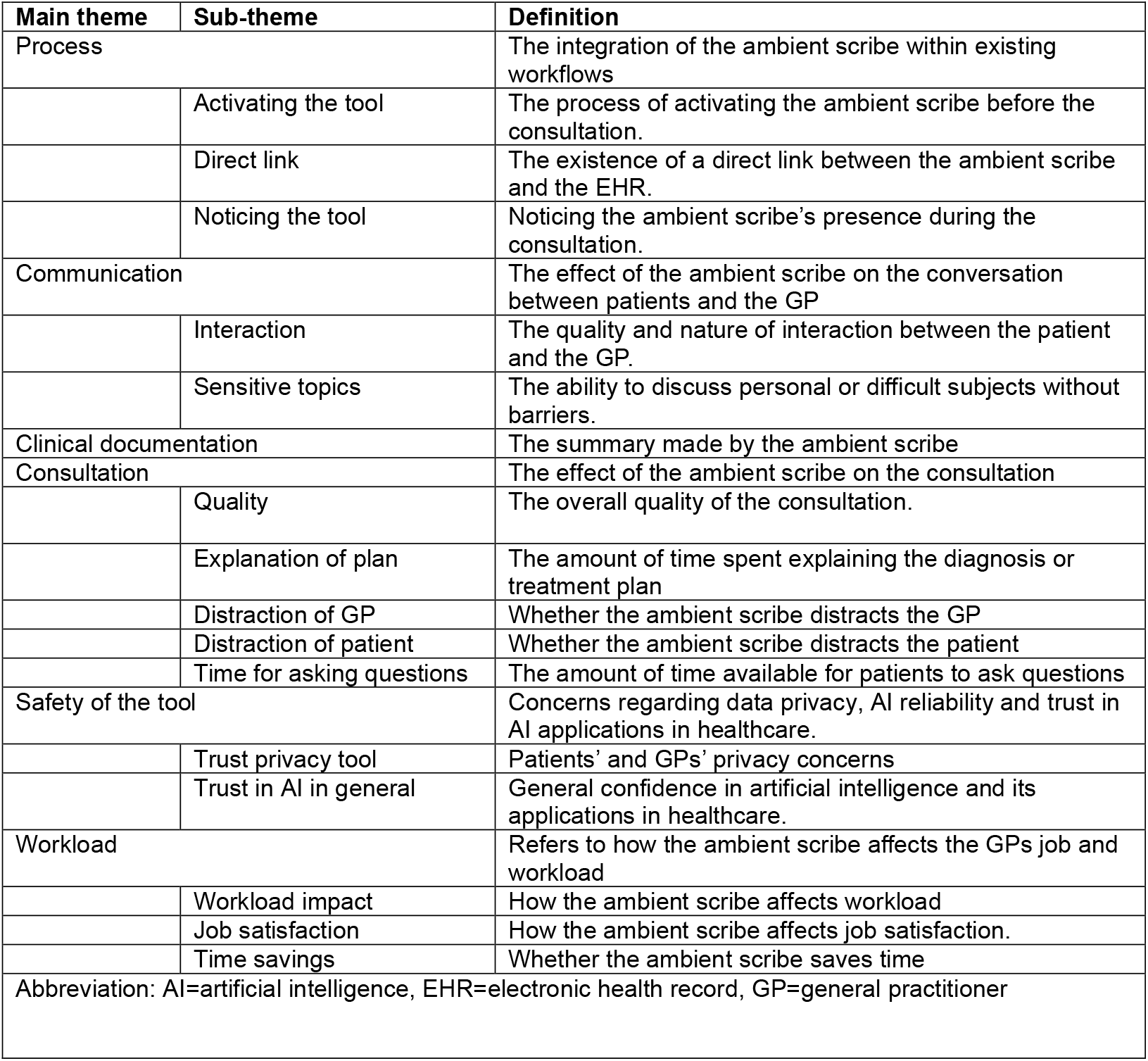
Themes, subthemes and definitions from the thematic analysis of interviews with patients and GPs.

| Main theme | Sub-theme | Definition |
| --- | --- | --- |
| Process |  | The integration of the ambient scribe within existing workflows |
|  | Activating the tool | The process of activating the ambient scribe before the consultation. |
|  | Direct link | The existence of a direct link between the ambient scribe and the EHR. |
|  | Noticing the tool | Noticing the ambient scribe's presence during the consultation. |
| Communication |  | The effect of the ambient scribe on the conversation between patients and the GP |
|  | Interaction | The quality and nature of interaction between the patient and the GP. |
|  | Sensitive topics | The ability to discuss personal or difficult subjects without barriers. |
| Clinical documentation |  | The summary made by the ambient scribe |
| Consultation |  | The effect of the ambient scribe on the consultation |
|  | Quality | The overall quality of the consultation. |
|  | Explanation of plan | The amount of time spent explaining the diagnosis or treatment plan |
|  | Distraction of GP | Whether the ambient scribe distracts the GP |
|  | Distraction of patient | Whether the ambient scribe distracts the patient |
|  | Time for asking questions | The amount of time available for patients to ask questions |
| Safety of the tool |  | Concerns regarding data privacy, AI reliability and trust in AI applications in healthcare. |
|  | Trust privacy tool | Patients' and GPs' privacy concerns |
|  | Trust in AI in general | General confidence in artificial intelligence and its applications in healthcare. |
| Workload |  | Refers to how the ambient scribe affects the GPs job and workload |
|  | Workload impact | How the ambient scribe affects workload |
|  | Job satisfaction | How the ambient scribe affects job satisfaction. |
|  | Time savings | Whether the ambient scribe saves time |
| Abbreviation: AI=artificial intelligence, EHR=electronic health record, GP=general practitioner |  |  |

### Process of using the tool

Several GPs reported that, because the tool was not directly integrated into their EHR, they occasionally forgot to activate the tool before the consultation started, particularly in urgent or complex cases. The need to manually copy and paste the generated summaries into the EHR was generally perceived as a minor inconvenience, requiring little additional time. However, GPs with lower levels of digital proficiency found this more burdensome. GPs using the integrated version also reported limitations. These were the inability to automatically transfer measurement data to the vital signs section and to produce separate summaries for consultations with multiple complaints.

Most patients did not notice the presence of the microphone or the use of the tool during consultations. Similarly, most GPs reported that patients were generally unaware of the tool and microphone, with almost none making any remarks about its presence.

### Communication

Some GPs suggested that less typing during consultations could enhance patient interaction, reduce screen time, and foster a more continuous connection with the patient through increased eye contact. One GP also noted that using the tool might help ensure that subtle patient cues or interactions are less likely to be missed. However, almost all GPs reported that they barely typed during consultations and thus perceived no difference in their consultation dynamics or patient interaction.

Most patients reported no difference in interaction compared with previous consultations, noting that GPs maintained attention, eye contact, and communication as usual. This was largely attributed to the fact that GPs already devoted sufficient attention and time to their patients during consultations. However, some patients felt they received more attention from their GP, with a few noticing increased eye contact during the consultation. They described this as contributing to a better conversation, making it easier to reach the core of their concerns and fostering a stronger sense of connection and trust in the GP.

Most GPs and patients noted that the tool could potentially pose a barrier for patients, particularly during consultations involving sensitive topics such as domestic violence, relationship problems, addiction, STDs or mental health issues. A few patients mentioned that they already found it difficult to talk about psychological problems and that the presence of the tool might slightly increase this barrier. GPs emphasized that they could easily deactivate the tool when a patient felt uncomfortable. When patients were directly asked whether they personally felt inhibited in discussing such topics while the tool was active, most reported that they did not experience any barriers. They indicated that they would feel comfortable discussing their complaints even if the tool was used in future consultations. Although several GPs and patients expected that older adults or people from different cultural backgrounds might experience greater discomfort, we observed no differences in the interviews between age groups, ethnic backgrounds, or education levels.

### Clinical documentation

Most GPs acknowledged that the AI-generated summaries were not always completely accurate, requiring them to review and adjust nearly every summary to ensure there were no errors. Several GPs reported that essential information or risk factors were occasionally missing, and one GP continued to make personal notes because he could not rely on the system. Almost all GPs mentioned that the physical exam was not correctly summarized by the ambient scribe tool, requiring them to manually type this section. The overall quality of the summaries tended to be lower for follow-up consultations, consultations involving more persons, phone calls, patient who switched between languages, or cases in which multiple complaints were discussed. However, other GPs noted that the tool was able to distinguish multiple complaints effectively in some situations. Overall, GPs noted that it was convenient to have most of the summary already written down, and they only needed to make the necessary adjustments. Some GPs found the AI-generated summaries particularly valuable for longer or more complex consultations, such as those with persistent somatic symptoms or psychiatric complaints, whereas others considered them more accurate and useful for shorter and simpler consultations. Nonetheless, many GPs found the ability to adjust the AI-generated summary using different templates very useful, for example by modifying the reason for encounter or refining the medical terminology.

Some patients suggested that using an ambient scribe tool could improve clinical documentation by making it more detailed, objective, and less influenced by the GP’s interpretation. Other patients expressed concerns that the summary might not capture the full essence of the conversation. Since the tool does not understand non-verbal interaction, it may miss nuances such as sarcasm or facial expressions. Nevertheless, other patients pointed out that GPs review and edit the notes before entering them in the EHR and therefore believed this would not affect the overall quality of the documentation.

### Consultation

Most patients thought the tool could give GPs more time to explain the diagnosis or treatment but did not believe it enabled them to ask more questions. When specifically asked, many reported no difference compared with previous consultations, as they felt there was already sufficient time for explanation and questions. Similarly, GPs stated that they always tried to take enough time to explain the diagnosis or treatment plan, with no noticeable difference when using the ambient scribe. A few patients believed that any time savings mainly occurred outside direct patient contact, allowing GPs to handle more administrative tasks, see additional patients, or address more complaints within the same consultation.

Almost all GPs reported that the tool did not cause any distraction during the consultation, with only a few exceptions such as when the wrong language was selected. All patients confirmed that they did not notice the GP being distracted by the tool. Both GPs and patients generally agreed that the tool did not distract the patients, as most were unaware of its use.

Some GPs also mentioned that using the tool could enhance the overall quality of the consultation. They explained that it helped them think in a more structured way, and the generated summary served as both a memory aid and as means to verify with patients whether the information had been recorded correctly. It also ensured that consultations were documented immediately, preventing delays in completing notes. However, one GP noted that summarizing was part of their clinical reasoning process, and that this could be lost when using the tool. Another GP felt that the quality of the consultation decreased, as the summaries became less accurate when the conversation deviated from the main topic or included small talk, making the GP less likely to engage in such interactions.

### Safety/privacy of the tool

Some patients and GPs expressed concerns about the safety of the tool, particularly regarding whether the transcripts used to generate the summaries were deleted immediately, where they were stored, and how securely they were protected. They were specifically worried about the potential risk of data breaches. However, most patients and GPs were not particularly worried about data security, with many patients expressing trust in their GP and confidence that confidential information is well protected within the Dutch healthcare system.

### Workload

Some GPs mentioned that the tool helped reduce their workload, noting that it was less demanding and required less cognitive effort to adjust the AI-generated summary than to type it themselves. A few GPs added that if the summaries would be more accurate, the tool could further improve efficiency. Most patients thought that the tool could reduce the workload for GPs. Some GPs did experience an increase in work satisfaction, noting that not having to do clinical documentation made consultations easier. Other GPs appreciated trying out new technologies, but the tool did not contribute to an increase in work satisfaction.

Most GPs experienced spending less time on clinical documentation and were running less behind on their consultations. One GP mentioned that the additional time gained was used to address more complaints within a single consultation, while others used it for practice related tasks. However, one GP found that the tool occasionally increased workload, particularly when seeing patients briefly between scheduled appointments or when additional time was required to reread and correct the summary. A few GPs also noted that, as they normally typed while patients were getting undressed for the physical exam, they now spent that time waiting, which felt less efficient. The tool was further described as inconvenient when making a referral, as the summary had to be completed while the treatment plan was often still being discussed. Some GPs mentioned they usually did not explain the tool to patients, as doing so would also take additional time. Almost all patients could imagine that the use of the ambient scribe tool for clinical documentation would save time, though some mentioned that correcting the summary could be time-consuming.

## Discussion

In this prospective, multicentre, longitudinal before–after mixed-methods study, we found that the use of an ambient scribe reduced documentation time in general practice. However, it did not affect total consultation time or patient experience. During the intervention period, documentation length increased, with more signs and plans recorded but fewer symptoms. The tool was applied in most consultations, except when technical issues arose, the GP forgot to activate it, or an intervention was performed. While the ambient scribe reduced GP workload, neither patients nor GPs perceived improvements in communication. The generated summaries were not always fully accurate, often missed non-verbal cues, and required GP review in all cases.

The research on ambient scribes has rapidly increased with the widespread use of LLMs(12-16, 18-24). However, most studies rely on aggregated retrospective data or surveys to measure time outcomes and do not adjust for differences between treatment and control period. In contrast, we used continuous external observation, the gold standard for measuring time outcomes, and demonstrated that an ambient scribe reduced documentation time by 42.7 seconds per consultation. This finding remained robust in both per-protocol and (post-hoc) sensitivity analyses, which adjusted for total documentation time and reason for consultation.(17) Our study, using gold standard methodology and adjusted analyses, confirms the results of prior studies reporting a reduction in documentation time(12-16, 23)

No difference in total consultation time was observed in the intention-to-treat analysis, with the effect estimate moving closer to zero in both the per-protocol and the post-hoc sensitivity analysis adjusted for reason for consultation. A possible explanation is that reductions in documentation time were offset by GPs spending more time on other parts of the consultation, for example by expanding the medical history or taking more time to explain the diagnosis and treatment. We also observed that GPs used a period of downtime during consultations to type the summary, which may have diminished the ambient scribe’s effect on reducing total consultation time. A different explanation is that, while reduced documentation time may shorten the overall consultation, the high variance in total consultation time could have limited the statistical power to detect such an effect. Future studies should base their sample size calculations on total consultation time and examine how the use of an ambient scribe influences different components of the consultation to better understand its impact on time outcomes.

In interviews, GPs mostly described the reduction in documentation time as leading to lower workload and improved work satisfaction, helping them feel less tired and less behind on consultations. These observations are consistent with previous research showing that ambient scribes can reduce burnout, cognitive burden, and task load.(12, 14-16, 20, 21, 23, 24) Considering both our quantitative and qualitative findings alongside prior studies, ambient scribes appear most effective at reducing workload and enhancing work satisfaction for healthcare providers. They are less effective at shortening consultation time or enabling providers to see more patients in a day.

Our study indicates that the use of an ambient scribe can have both positive and negative effects on patient–doctor communication. On one hand, some patients reported that the tool enhanced their sense of connection and trust, citing increased eye contact and better conversation. In contrast, most GPs did not perceive an improvement, noting that they already typed minimally during conversations, typically using idle moments for documentation. Previous studies, mostly from the healthcare provider’s perspective, have shown mixed evidence on this topic, with some reporting improved communication and others finding no effect.(12, 14, 19, 20, 22, 24) Future research should use more objective measures to assess whether ambient scribes can mitigate the impact of the EHR on communication.(40)

Conversely, some patients felt that the presence of an ambient scribe could create a barrier when discussing sensitive issues. Although most participants did not feel inhibited, there may be a small group of patients that are impacted. Our data showed no differences across age, background, or education. However, prior research suggests that digital tools may exacerbate health disparities, underscoring the need to ensure that the implementation of ambient scribes does not compromise accessibility of care for these subgroups.(41)

When comparing notes generated by the ambient scribe with those written by the GP, we observed that the notes of the ambient scribe were generally longer across most sections. However, they contained more substantive information only in the signs and plan sections. At the same time, fewer symptom variables were recorded. Interviews revealed that this occurred because the ambient scribe often failed to correctly summarise the physical examination, requiring the GP to add this information manually. Yet, even with these additions, fewer symptom variables were ultimately documented.

A possible explanation is that, because the scribe already produces summaries for the other sections, GPs may invest less effort in completing the physical examination notes than they would if they were writing the entire record themselves. This may create overreliance on the scribe, with insufficient correction by the GP, reducing the quality of documentation for this part of the consultation. This risk of degraded documentation quality warrants further study.

The problem may be compounded by the fact that ambient scribes occasionally hallucinate, requiring GPs to carefully review and adjust each summary, an issue also reported in earlier studies (14, 21, 24, 25, 45). Patients likewise expressed concerns that the scribe might miss sarcasm, non-verbal cues, or unspoken reflections from the GP. Future research should therefore assess documentation quality not only by comparing notes with consultation transcripts, but also by examining their practical value both for healthcare providers who use them in follow-up care and for patients when reading back their own records.

Ambient scribes may have mixed effects on the clinical reasoning process. Some GPs perceived an improvement, noting that the scribe functioned as a memory aid and allowed them to verify information directly with the patient. However, one GP expressed concern that delegating note-taking might remove an important moment of reflection that typically occurs when clinicians summarise the consultation themselves. Given that there is an important role of summarisation in the clinical reasoning process, future research should explore this potential unintended consequence.(42)

The main strength of our study is that, to our knowledge, it is the first prospective mixed-methods study to use real-time outcome measurements on ambient scribing, ensuring robust results. Although our study focused on general practitioners, the results are likely transferable to other outpatient settings, including medical specialty care as the findings are consistent with previous research on ambient scribing in other contexts.(12-24) However, our results extend the evidence base by providing more precise data on time-related outcomes and by offering a deeper understanding of potential adverse effects, including possible impacts on clinical reasoning, loss of detail regarding symptoms and measurements in clinical summaries, and patients’ perspectives on ambient scribing.

Some limitations should be acknowledged. First, participation was limited to GPs who were willing to adopt ambient scribing. These GPs may have held more favourable attitudes toward ambient scribes, potentially influencing both their experiences and overall usage rates. Second, most participating practices served relatively few patients from deprived areas, and the majority of interviewed patients had higher education levels and a Western background. Individuals with lower SES and diverse migration backgrounds were underrepresented. Prior research indicates that patients from lower SES groups may show greater distrust toward AI in healthcare, often linked to concerns about data privacy or limited familiarity with the technology.(43) This underrepresentation may reduce generalizability of our findings. Third, the GPs in our sample scored highly on both digital skills and UTAUT questionnaires and were relatively young. Our results may not fully capture the perspectives of older GPs or those less experienced with digital tools. Future research should make a concerted effort to include these underrepresented groups and conduct subgroup analyses to evaluate the effectiveness and acceptability of ambient scribing across different populations. This is particularly important given the risk that digital innovations may inadvertently widen existing health disparities.(46)

To increase the impact of ambient scribes in healthcare, developers should prioritize the following areas. First, integration with the EHR to minimize the time spent copying and pasting summaries. Second, improving the accuracy and quality of the generated summaries, particularly regarding symptoms and measurements, to further reduce the time clinicians spend on summarization and to prevent less detailed documentation of these variables. Third, adding advanced functionalities based on the content of the summary, such as drafting a referral or ordering diagnostic tests, to lessen the administrative burden on healthcare providers.

Concluding, ambient scribing in general practice reduces documentation time and workload, improves job satisfaction, and potentially patient–provider communication. However, it also results in less detailed documentation of clinical signs in the EHR, does not increase patient throughput, could reduce accessibility for patients with sensitive issues, and it may interfere with aspects of the clinical reasoning process.

## Patient and public involvement

Rijnmond Dokters, the regional care group of GPs in Rotterdam, the Netherlands, contributed to the study by providing expertise in general practice and digital innovation as well as partial funding. Their input helped shape the study design, interpret the data, and write the report. This involvement was led by their chief medical information officer and practicing GP (LJ), who is also a co-author of this report

## Ethical approval

The medical ethical review committee of the Erasmus MC deemed the study to not be subject to the Medical Research Involving Human Subjects Act and provider a waiver (MEC-2024-0286).

## Data availability

The data supporting the findings of this study are available from the corresponding author (RvL) upon reasonable request. Due to privacy considerations, the data are not publicly accessible. Metadata are available, and data requests can be submitted via the DataVerseNL repository (https://doi.org/10.34894/ITBO7X). Syntax for the analyses can be found on GitHub (https://github.com/reiniervlinschoten/ai_scribe).

## Contributors

RvL and LJ contributed to the conception and design of the study. RvL and CvL contributed to participant inclusion and data collection and were responsible for project administration. RvL and EB were responsible for project supervision. RvL and CvL accessed and verified the underlying data. RvL was responsible for statistical analysis. All authors contributed to interpreting the data. CvL and RvL wrote the first draft of the manuscript. All authors reviewed and approved the manuscript for submission. All authors had access to all data and shared final responsibility for the decision to submit for publication.

