## Supplemental File 1 for "Ambient scribe in general practice: a multi-perspective before-after longitudinal mixed-methods study"

### Supplementary Methods

**Training and validity of outcome assessors**
Research assistants, trained to ensure consistency and accuracy, collected all data. The same assistants conducted baseline and intervention measurements for each GP to minimize inter-observer variability. Training included measurement and review of consultation videos to standardize task categorization for time outcomes. Assessment of documentation quality was similarly trained through review of sample notes. Last, all assistants were trained in interview techniques with mock interviews and were given the topic guide to guide the semi-structured interview. Interviewers had no experience with the ambient scribe and were not (previously) acquainted with the company, any of the GPs, or the patients involved in the study.

**Topic guide**
This is the interview guide for the semi-structured interviews for the study *"AI Transcription and Reporting in General Practice: A Longitudinal Study."* The interviews will last approximately 10-20 minutes.
Participants will be approached with the question whether they are willing to participate in a face-to-face interview or an interview done with MS Teams or a phone call, based on:

- Having participated in a consultation as a **patient** where Juvoly QuickConsult was used
- Having used Juvoly QuickConsult as a **general practitioner**

*Objective*
The aim of the interview is to explore how the GP/patient experiences the use of Juvoly QuickConsult, whether the system improves the consultation, and/or reduces workload.

*Preparation*
Discuss before starting the interview:

- Introduce the interviewer and explain the study/research goal
- Emphasize that participation is voluntary and participants may stop at any time
- Ask for consent to record the interview
- Start the recording in MS Teams and begin by stating the date and participant number

"Your responses will be processed and analysed anonymously, so they cannot be traced back to you. That's why we are recording the interview, also anonymously. There are no right or wrong answers in this conversation. You may stop the interview at any time."

*General interview tips*

- Follow up to explore what's behind the participant's answers: "Can you tell me more about that?", "In what way?", "How come?", "Can you give an example?"
- Try to empathize with the participant's situation to ensure a natural flow of conversation.
- Ask open-ended questions
- Avoid leading questions
- Do not express your own opinion or judgment

The questions marked with a dash (-) are useful for exploring a topic. Let the GP/patient speak as much as possible. Once they've answered, check if any bullet points haven't been mentioned, and ask about those specifically.

*Questions for the GP*

- What do you think of Juvoly QuickConsult?
- Does the system help you? How?
    - Workload? Focus? Job satisfaction?
- Does the system help the patient? How?
    - More time to explain or ask questions?
- Are there any disadvantages of the system for you? Which ones?
    - Distraction?
- Are there any disadvantages of the system for the patient?
    - Distraction? No desire to talk about certain topics / share certain information? Less opportunity to ask questions?

*Questions for the patient*

- General information: Age, highest completed education, proficiency in Dutch (poor, average, good), country of birth, country of birth of father/mother, chronic illnesses
- How did you experience the consultation?
- Have you noticed that the tool was used during the consultation?

- What do you think of the system the doctor used to record the consultation?
- Does the system help the doctor? How?
  - More time for explanation?
- Does the system help you? How?
  - More time for clarification of the diagnosis/treatment of plan?
- Are there any disadvantages of the system for the doctor? How?
  - Distraction for the doctor?
- Are there any disadvantages of the system for you?
  - Distraction? Feeling a threshold to talk about certain topics / share certain information? Less opportunity to ask questions?

"Thank you very much for your participation."

#### Supplementary Tables

| Task Category | Description |
|---|---|
| **Communication** | |
| Explaining tool | Explaining the ambient scribe to the patient |
| Medical History | Taking the medical history and exploring complaints of the patient |
| Plan | Explaining the diagnosis or the treatment plan |
| Colleague | Consulting a colleague |
| Small talk | Small talk with patient or family |
| Other patient | Actions concerning another patient |
| **Hands-on** | |
| Physical exam | Conducting the physical examination |
| Documentation | Writing clinical documentation or correcting the AI-generated summary |
| Waiting on the ambient scribe | Waiting for the ambient scribe to process the consultation |
| Administration | Doing administrative duties (e.g., prescriptions, referrals) |
| Copy/pasting | Copying and pasting the AI-generated summary from the website into the electronic health record |
| Intervention | Performing an intervention (e.g., vaccinations, biopsies) |

**Supplementary Table 1.** Definitions of work activities and tasks

| Characteristic[1] | Baseline<br>N = 264 | Intervention<br>N = 271 |
|---|---|---|
| Patient age (years) | 49 (25, 67) | 53 (32, 69) |
| Female | 155 (59%) | 172 (63%) |
| Number of complaints | 1 (1, 1) | 1 (1, 1) |
| Planned consultation time | | |
| 10 minutes | 10 (3.8%) | 12 (4.4%) |
| 15 minutes | 241 (91%) | 242 (89%) |
| 20 minutes | 6 (2.3%) | 10 (3.7%) |
| 30 minutes | 7 (2.7%) | 7 (2.6%) |
| Tool used | 0 (0%) | 244 (90%) |
| Reason tool not used | | |
| Forgotten | - | 7 (26%) |
| GP decision | - | 1 (3.7%) |
| Intervention | - | 5 (19%) |
| Technical difficulties | - | 13 (48%) |
| Wrong language | - | 1 (3.7%) |
| Surveys consented/sent | 208 (79%) | 138 (51%) |
| Surveys answered | 120 (58%) | 71 (51%) |

[1] Median (Q1, Q3); n (%)

**Supplementary Table 2.** Characteristics of consultations

| Characteristic | Baseline[1]<br>N = 291 | Intervention[1]<br>N = 291 | Difference[2] |
|---|---|---|---|
| ICPC code | | | |
| Circulatory tract | 21 (6.3%) | 26 (7.7%) | 1.4% |

| | | | |
|---|---|---|---|
| Digestive tract | 21 (6.3%) | 20 (5.9%) | -0.4% |
| Ear | 24 (7.2%) | 16 (4.7%) | -2.5% |
| Endocrine tract/metabolism | 8 (2.4%) | 13 (3.8%) | 1.4% |
| Eye | 9 (2.7%) | 12 (3.5%) | 0.8% |
| Female genitalia and breasts | 13 (3.9%) | 8 (2.4%) | -1.5% |
| General/non-specified | 32 (9.6%) | 17 (5.0%) | -4.6% |
| Haematological | 1 (0.3%) | 2 (0.6%) | 0.3% |
| Male genitalia and breasts | 3 (0.9%) | 6 (1.8%) | 0.9% |
| Musculoskeletal system | 54 (16%) | 56 (17%) | 1% |
| Nervous system | 10 (3.0%) | 21 (6.2%) | 3.2% |
| Pregnancy/birth/contraception | 7 (2.1%) | 1 (0.3%) | -1.8% |
| Psychological problems | 20 (6.0%) | 26 (7.7%) | 1.7% |
| Respiratory tract | 49 (15%) | 44 (13%) | -2% |
| Skin | 49 (15%) | 58 (17%) | 2% |
| Social problems | 7 (2.1%) | 5 (1.5%) | -0.6% |
| Urinary tract | 5 (1.5%) | 8 (2.4%) | 0.9% |

[1] n (%)
[2] Absolute difference in percentage points between periods
Abbreviation: ICPC=International Classification of Primary Care
**Supplementary Table 3.** Complaints discussed during consultations as categorised per ICPC code

| **Characteristic** | **Baseline** N = 264[1] | **Intervention** N = 271[1] |
|---|---|---|
| Total consultation time | 901 (684, 1,248) | 861 (625, 1,164) |
| Time spent explaining toole | 0 (0, 0) | 0 (0, 13) |
| Time spent taking the medical history | 224 (134, 362) | 208 (111, 380) |
| Time spent doing the physical examination | 118 (45, 180) | 101 (32, 182) |
| Time spent on explaining diagnosis and treatment | 212 (136, 299) | 229 (137, 321) |
| Time spent consulting a colleague | 0 (0, 0) | 0 (0, 0) |
| Time spent on other tasks | 0 (0, 0) | 0 (0, 0) |
| Time spent on small talk | 0 (0, 29) | 0 (0, 11) |
| Time spent on typing documentation | 138 (95, 207) | 81 (48, 136) |
| Time spent on administrative duties | 76 (26, 132) | 70 (21, 143) |
| Time spent doing an intervention | 0 (0, 0) | 0 (0, 0) |
| Time spent waiting on the ambient scribe | 0 (0, 0) | 4.0 (0, 7) |
| Time spent copy and pasting the summary | 0 (0, 0) | 11 (0, 21) |
| Time spent multitasking | 31 (7, 79) | 17 (3, 50) |
| Time spent on the primary outcome: typing documentation, waiting on the scribe and copy and pasting the summary | 138 (95, 207) | 102 (68, 165) |

[1] Median seconds (Q1, Q3)
**Supplementary Table 4.** Distribution of time outcome variables across the baseline and intervention periods

| **Name** | **Estimate** | **95% CI** | **p-value** |
|---|---|---|---|
| **Time outcomes[1]** | | | |
| Documentation time | -41.00 | [-53.47; -29.95] | <0.0001 |
| **Note length[2]** | | | |
| Subjective | 1.17 | [1.07; 1.29] | 0.0006 |
| Objective | 0.87 | [0.71; 1.04] | 0.127 |
| Assessment | 1.18 | [1.01; 1.38] | 0.038 |
| Plan | 1.60 | [1.44; 1.8] | <0.0001 |
| **Number of variables in note[2]** | | | |
| Signs | 1.12 | [1.01; 1.24] | 0.031 |
| Context | 1.06 | [0.92; 1.21] | 0.369 |
| Symptoms | 0.80 | [0.68; 0.93] | 0.004 |
| Diagnosis | 0.98 | [0.84; 1.14] | 0.773 |
| Plan | 1.19 | [1.08; 1.3] | 0.0003 |

| |
|---|
| Abbreviation: CI=confidence interval<br>[1] Difference on an additive scale<br>[2] Difference on a multiplicative scale<br>**Supplementary Table 5.** Parameter estimates, 95% CIs and p-values of the sensitivity analyses adjusted for non-documentation time |

| **Name** | **Estimate** | **95% CI** | **p-value** |
|---|---|---|---|
| **Time outcomes**[1] | | | |
| Documentation time | -42.84 | [-54.95; -30.62] | <0.0001 |
| Consultation time | -47.70 | [-110.83; 15.61] | 0.164 |
| **Note length**[2] | | | |
| Subjective | 1.22 | [1.11; 1.33] | <0.0001 |
| Objective | 0.88 | [0.73; 1.05] | 0.166 |
| Assessment | 1.18 | [1.01; 1.38] | 0.048 |
| Plan | 1.66 | [1.47; 1.86] | <0.0001 |
| **Number of variables in note**[2] | | | |
| Signs | 1.17 | [1.05; 1.28] | 0.005 |
| Context | 1.15 | [0.95; 1.26] | 0.187 |
| Symptoms | 0.76 | [0.68; 0.94] | <0.0001 |
| Diagnosis | 0.97 | [0.83; 1.16] | 0.827 |
| Plan | 1.17 | [1.11; 1.36] | <0.0001 |

Abbreviation: CI=confidence interval
[1] Difference on an additive scale
[2] Difference on a multiplicative scale
**Supplementary Table 6.** Parameter estimates, 95% CIs and p-values of the per protocol analyses

| Name | Estimate | 95% CI | p-value |
|---|---|---|---|
| Time outcomes[1] | | | |
| Documentation time | -42.16 | [-54.22; -29.82] | <0.0001 |
| Consultation time | -45.38 | [-107.39; 18.42] | 0.162 |

Abbreviation: ICPC=International Classification of Primary Care, CI=confidence interval
[1] Difference on an additive scale
**Supplementary Table 7.** Parameter estimates, 95% CIs and p-values of the post-hoc sensitivity analysis adjusted for ICPC code

| **Characteristic** | **N = 48**[1] |
|---|---|
| Age (years) | 64 (56, 72) |
| Female | 27 (56%) |
| Education level | |
| Low | 19 (40%) |
| Middle | 11 (23%) |
| High | 18 (38%) |
| Dutch proficiency | |
| Poor | 1 (2.1%) |
| Fair | 3 (6.3%) |
| Good | 44 (92%) |
| Migration background | |
| Dutch background | 40 (83%) |
| First generation non-western migration background | 3 (6.3%) |
| First generation western migration background | 2 (4.2%) |
| Second generation non-western migration background | 1 (2.1%) |
| Second generation western migration background | 2 (4.2%) |
| Self-reported chronic disease | 19 (40%) |

[1] Median (Q1, Q3); n (%)

**Supplementary Table 8.** Demographic characteristics of interviewed patients
